# DIPHYSIO study protocol: a pilot multi-centre open-label randomised controlled trial assessing prevention of recurrent DIverticulitis through the use of pelvic floor PHYSIOtherapy

**DOI:** 10.64898/2025.12.28.25343115

**Authors:** Muhammad Imran Aumeerally, Christopher Gillespie, Andrea Warwick, Allison Bryant, Kate Hooper, Ferdinand Ong, Matthew Burstow, Melanie Walkenhorst

## Abstract

**Introduction:** Despite the ubiquity of diverticular disease, the options for reducing the risk of recurrent diverticulitis remain limited and the pathogenesis remains incompletely understood. While high intraluminal pressures within the distal colon and rectum have been proposed as a possible association with diverticular disease, studies on this relationship have been few, inconsistent and not generalisable. The investigators of this pilot study propose that the repeated transient high intraluminal pressures generated within the distal colon and rectum due to ineffective defecatory technique may predispose some patients to an increased risk of diverticulosis and diverticulitis. Therefore, by correcting defecatory technique through the implementation of pelvic floor physiotherapy (PFPT), the investigators hypothesise that there would be a reduction in the risk of recurrent diverticulitis.

**Methods and analysis:** This pilot multi-centre open-label randomised controlled trial will be conducted at Queen Elizabeth II Jubilee Hospital (QEII) and Logan Hospital (LGH) in Brisbane, Queensland, Australia. Eligible adult patients admitted with acute diverticulitis will be considered for enrolment and randomised into two groups in a 1:1 allocation ratio. The aim is to recruit 40 patients with 20 patients per group. The control group will receive standard of care dietary advice. The intervention group will receive PFPT as an outpatient within 4 weeks of discharge. The primary endpoint will be the risk of readmission with recurrent diverticulitis within a 12-month follow-up period. Secondary endpoints will be the risk of surgical intervention and/or interventional radiology (IR) procedure in the subgroup of patients readmitted with recurrent diverticulitis. Feasibility outcomes will review patient compliance and completeness of data collection. Results of this trial will inform study design and sample size required in a larger prospective study.

**Ethics and dissemination:** Approval was obtained from the Human Research Ethics Committee at the participating centre. Results will be submitted for publication in a peer-reviewed journal.

**Trial registration number:** ACTRN126250009274426.

**STRENGTHS AND LIMITATIONS OF THIS STUDY:**

- This pilot RCT is the first prospective study to assess the correction of defecatory dysfunction as a method for reducing the risk of recurrent diverticulitis
- A practical design with ease of reproducibility that will inform a larger study adequately powered for hypothesis testing
- Open-label design poses risk of performance bias
- Lack of standardisation for pelvic floor physiotherapy interventions may impact generalisability outside of facilities with dedicated pelvic floor units

## BACKGROUND

Colonic diverticular disease is a common condition that describes the development of asymptomatic diverticula and the subsequent complications including inflammation, perforation and bleeding. Diverticula are the herniations of mucosa and submucosa through the muscular layer of the colon typically located either side of the taenia coli where the vasa recta penetrate the colonic wall. Inflammation of the diverticula is known as diverticulitis and is typically characterised by left lower quadrant abdominal pain, altered bowel motions and pyrexia^1^.

Over time, the prevalence of diverticular disease in Australia and Western societies has markedly increased and appears to be trending towards younger patients with more complicated disease^2-6^. An estimated 5% to 25% of patients with diverticulosis are expected to develop diverticulitis at least once in their lifetime^5-7^. After an episode of acute diverticulitis, approximately 20% to 55% will suffer recurrent episodes of diverticulitis^4, 8, 9^. These patients with recurrent diverticulitis are at risk of repeat hospitalisation, need for invasive interventions, chronic pain, loss of productivity and reduced quality of life.

The pathogenesis of diverticular disease remains incompletely understood. The current evidence points to a multifactorial process both in the development of diverticulosis and diverticulitis with several overlapping risk factors shared^10^. These risk factors include dietary fibre intake, genetic predisposition, altered microbiota composition, colonic wall structural changes and colonic dysmotility.

Reduced dietary fibre intake has been proposed as a potential cause of diverticular disease since the 1970s^11^. This association has been observed in several prospective studies and further established in meta-analyses^12-14^. Conversely, the evidence for high intraluminal pressures due to colonic dysmotility has been mixed^15-17^. Increasingly the focus of many studies has shifted towards the role of the gut microbiota and the specific changes in the colonic wall structure which may cause dysregulation of normal peristalsis^18, 19^.

Dyssynergic defecation falls under the umbrella of defecatory dysfunction and is characterised by a failure of rectal propulsion, a failure of adequate anal sphincter relaxation or a combination of both^20^. The investigation team proposes that these repeated transient high pressures generated during defecation may contribute to the development of diverticulosis and diverticulitis in some individuals. Therefore, by correcting defecatory technique through PFPT, the investigators propose that a reduced risk of diverticulitis should be observed.

Given the lack of widely accepted measures to prevent diverticulitis, initiatives such as PFPT offer a potentially non-invasive and affordable opportunity to help reduce recurrent diverticulitis, the need for invasive interventions and the impact on patient quality of life.

## METHODS

### Hypothesis

The use of PFPT reduces the short-term risk of recurrent diverticulitis in patients compared to those who do not have PFPT.

### Objectives

The primary objective of the DIPHYSIO pilot study is to:

- Assess risk of readmission with recurrent diverticulitis in patients who receive PFPT and patients who do not receive PFPT following discharge from hospital for acute diverticulitis.

The secondary objectives of the DIPHYSIO pilot are to assess the subgroup of patients readmitted with recurrent diverticulitis and to:

- Assess risk of surgical intervention for recurrent diverticulitis in patients who receive PFPT and patients who do not receive PFPT following discharge from hospital for acute diverticulitis.
- Assess risk of IR procedure for recurrent diverticulitis in patients who receive PFPT and patients who do not receive PFPT following discharge from hospital for acute diverticulitis.
- Assess composite risk of surgery or IR procedure for recurrent diverticulitis in patients who receive PFPT and patients who do not receive PFPT following discharge from hospital for acute diverticulitis.

The feasibility objectives of the DIPHYSIO pilot study are to:

- Assess participant compliance and retention
- Assess completeness of data collection processes

### Study design

This study is a pilot multi-centre open-label randomised controlled trial at QEII and LGH located within the Metro South Health (MSH) service in Brisbane, Queensland, Australia. In line with the SPIRIT 2025 Statement, this study has been registered with the Australian New Zealand Clinical Trials Registry (ANZCTR) on the 29^th^ August 2025^*21*^. Given the involvement of human participants, this study will be compliant with the guidelines set out by the National Statement of Ethical Conduct in Human Research 2025 and the Declaration of Helsinki^22, 23^. Ethics approval has been granted by MSH Human Research Ethics Committee (HREC/2025/QMS/120559) on the 15^th^ September 2025. Research governance approval has been granted by MSH Research Governance Office (SSA/2025/QMS/120559) on the 29^th^ October 2025.

### Eligibility criteria

Adult patients (aged 18 years or older) admitted to QEII or LGH with acute diverticulitis affecting the left colon (descending colon or sigmoid colon) will be considered eligible for the study. Patients who require emergency surgery or IR procedure for diverticulitis prior to enrolment are not eligible. Patients with diverticulitis of the caecum, ascending colon or transverse colon will not be considered eligible. Additionally, patients who have been admitted electively for surgical management of diverticular disease are not eligible (*Figure 1*).

**Figure 1.**
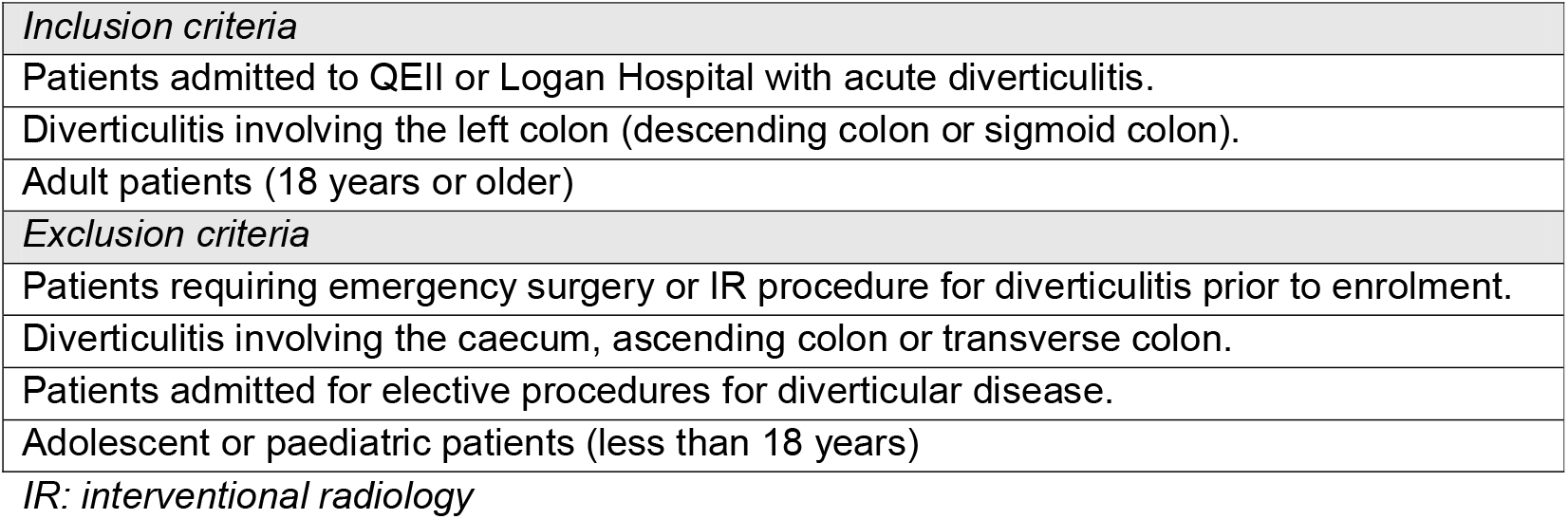
Eligibility criteria. IR: interventional radiology

### Control

Participants randomised into the control group will receive the usual standard of care following an episode of diverticulitis. This includes education on dietary fibre intake aiming for 25-30g fibre per day and oral fluid intake aiming for 8-10 glasses of water per day. No additional study related appointments or interventions will be required. Further review in clinic or investigations such as colonoscopy may be arranged at the discretion of the treating surgeon.

### Intervention

Participants randomised into the intervention group will receive PFPT in addition to standard care advice on fibre and fluid intake. Participants in the intervention group will be referred for a one-hour initial assessment with a pelvic floor physiotherapist in an outpatient setting within 3-4 weeks of discharge from hospital. The participant will receive an individualised assessment and tailored treatment program based on assessment findings. The tailored treatment program may include therapeutic exercises, manual therapy, instrument-based biofeedback, behavioural strategies and lifestyle modification strategies. Further PFPT appointments may be required every 3-4 weeks for a maximum of 6 months at the discretion of the physiotherapist. This is the standard of practice for PFPT assessments in MSH. Further review in clinic or investigations such as colonoscopy may be arranged at the discretion of the treating surgeon.

### Patient selection, recruitment, enrolment

Potential participants who meet eligibility criteria will be identified by the treating surgical teams of QEII and LGH. Potential participants will be approached by a member of the treating surgical team within 24 hours prior to expected discharge from hospital. The diagnosis of diverticulitis must be made radiologically or based on the clinical assessment of the treating surgeon. Consent will be preferentially obtained by a member of the treating surgical team for enrolment of the study. At the time of discharge from hospital, the participant will be enrolled in the study and will be randomly allocated by the lead investigator into either the control or intervention group. The randomisation key will be generated using the National Institute of Health (NIH) Clinical Trial Randomization Tool to allocate participants into equal groups with a maximum tolerated imbalance (MTI) of 3. Only the lead investigator will have access to the encrypted and password-protected randomisation key. The participant will be advised of the outcome of the randomisation.

### Patient and public involvement

Neither patients nor public individuals were involved in the conceptualisation or design of this study and will not be involved in the conduct, data collection, analysis, reporting or dissemination of this study.

### Data collection

Data collection and reporting will be performed in line with the SPIRIT 2025 Statement. Data monitoring will be conducted annually by MSH HREC as per local audit and review processes. In the event that an adverse event occurs in relation to data management, this will be reported to MSH HREC by the investigation team.

A linkage dataset file will contain reidentifiable data that links patients to their study ID. Only the lead investigator will have access to the linkage dataset file which will be encrypted and password-protected. Deidentified participant data will be collected in a separate encrypted and password-protected file with access granted only to members of the investigation team. Deidentified data will be stored for 15 years from the date of publication.

The data fields for collection will include participant demographics, the clinical course of diverticulitis prior to enrolment, and the outcomes (*Figure 2*).

**Figure 2.**
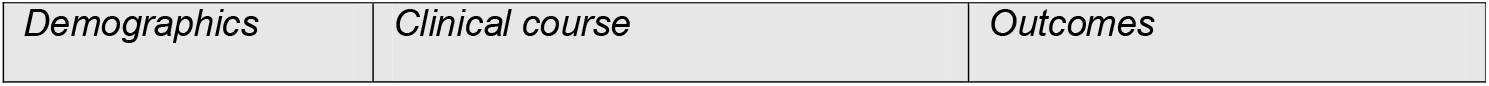
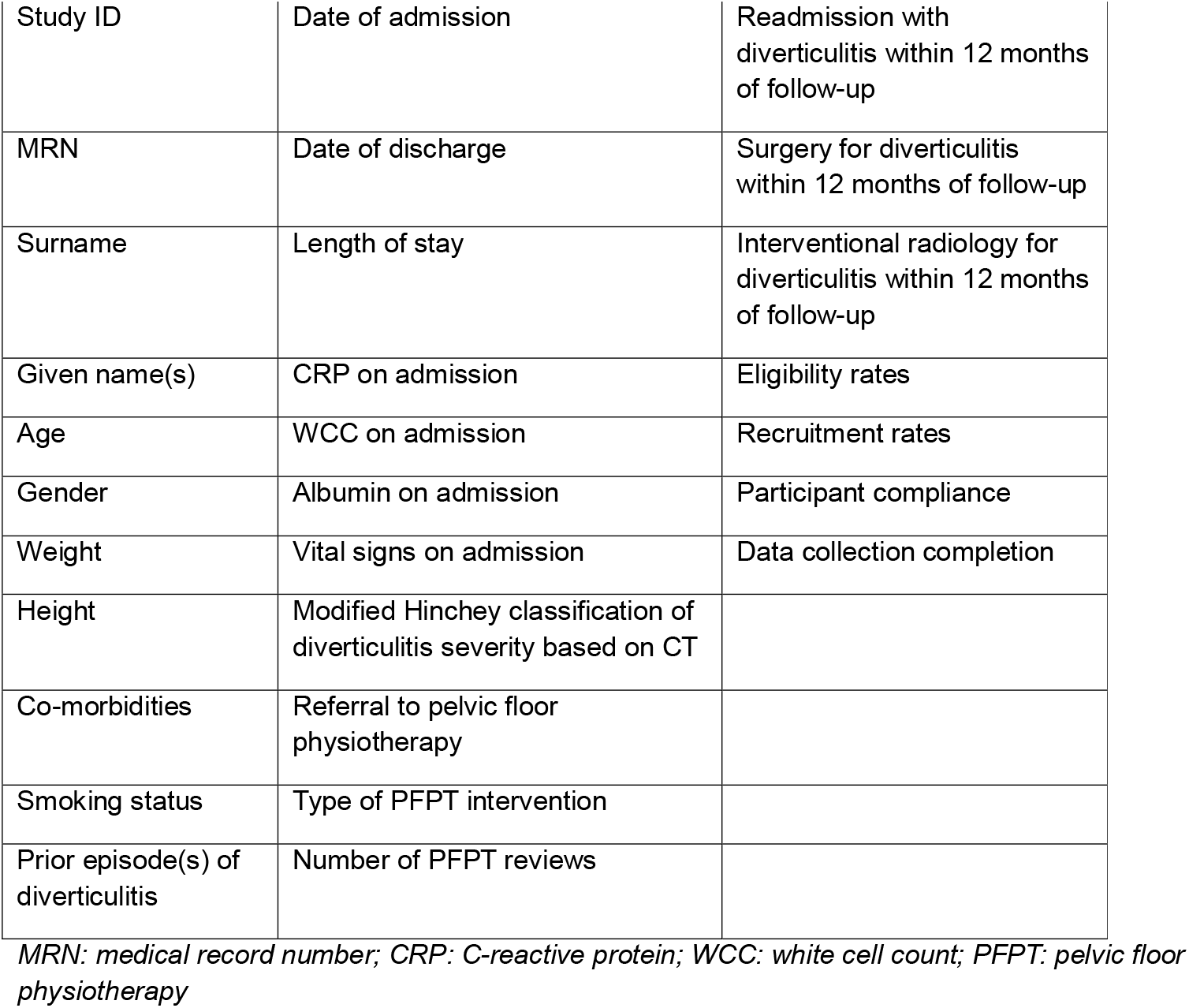
Data fields for collection. MRN: medical record number; CRP: C-reactive protein; WCC: white cell count; PFPT: pelvic floor physiotherapy

### Outcomes

The primary outcome for this pilot randomised controlled trial is the risk of readmission due to recurrent diverticulitis. The secondary outcomes for this study will be an assessment of the subgroup of participants who are readmitted with recurrent diverticulitis. In this subgroup, the investigators will assess the risk of requiring a surgical intervention for diverticulitis, the risk of requiring an IR procedure for diverticulitis and the composite risk of requiring either a surgical intervention or IR procedure for diverticulitis. The investigators will conduct a chart review of each participant every 2 months to check if the participant has been readmitted to hospital with recurrent diverticulitis and to check if a primary outcome or secondary outcome event has transpired. Feasibility will review participant compliance, withdrawal rates and data completeness of data collection. Issues around logistics (e.g. recruitment, storage of data, delays in time to PFPT review, etc.) will be routinely monitored by investigation team and published.

### Withdrawal

Participants may withdraw from the study at any time or may be withdrawn at the discretion of the treating surgeon. Participants may be withdrawn if the investigation team or ethics committee determine there are grounds to terminate the study. Participants who withdraw may consent to have their trial-related data expunged from the study.

### Statistical analysis

An a priori power analysis has not been performed as this is a pilot study. Statistical analysis will be conducted after all data points have been collected at the completion of the follow-up period.

Participants randomised to the PFPT intervention group will be evaluated using a modified intention-to-treat protocol. Participants who are allocated to PFPT and attend at least one PFPT clinic session will be included in the analysis. Participants who are allocated to PFPT and fail to attend at least one PFPT clinic session will be excluded from the study altogether.

Data will be analysed with the Stata statistical software package (Stata/SE 18.0 for Windows, StataCorp LLC, Texas, USA). Descriptive statistics will be used to compare baseline characteristics including demographics, comorbidities and clinical details of diverticulitis prior to enrolment. Appropriate inferential statistics will be used to compare primary and secondary outcomes. The publication of p-values will not be published as this is a pilot study. Instead, confidence intervals will be published to demonstrate the level of precision and certainty in the outcomes. Descriptive statistics will be used to demonstrate patient compliance and completeness of data collection.

## DISCUSSION

This study aims to evaluate the role of PFPT in the prevention of recurrent diverticulitis. The study is based on the proposition that an ineffective defecatory technique may predispose individuals to the development of diverticulosis and subsequently diverticulitis. It is proposed that the repeated transient increase in intraluminal pressure within the distal colon and rectum contribute to the structural changes in the colonic wall that are seen in diverticulosis.

Additionally, this high-pressured system may also result in an increased faecal turbulence resulting in obstruction of diverticula and therefore diverticulitis.

Studies on intraluminal pressures have been performed as far back as the 1960s and identified exaggerated pressure increases in patients with diverticulosis compared to those without diverticulosis after the gastrocolic reflex was elicited or a pharmacologic stimulant was used^15, 16^. A more contemporary study by Jaung et al used high-resolution manometry but did not observe a difference in pressures^17^. Further, Jaung et al conducted a systematic review and found the collated literature to be sparse in number and heterogenous in methodology^24^.

A forthcoming publication of a retrospective cohort study is the first to demonstrate a statistically significant reduction in recurrence rates of diverticulitis between patients referred for PFPT compared to those who were not^25^. The retrospective nature and small numbers of patients resulted in selection bias and limited ability to control confounders. Therefore, there was a need for the development of a randomised controlled trial to corroborate these findings.

If this practical pilot study demonstrates feasibility, the investigators will proceed with a larger randomised controlled trial that is adequately powered to detect a clinically significant difference in the risk of recurrent diverticulitis between patients who have PFPT and those who do not. The potential benefit of this intervention is that it is non-invasive and affordable to implement. It would also add to the limited existing options for prevention of recurrent diverticulitis and potentially reduce the risk of surgical intervention in patients with diverticular disease.

Due to the open-label design of this study, there is an inherent risk of performance bias. While it would be impractical to blind the participant or the physiotherapist, the treating surgeon who makes the diagnosis of recurrent diverticulitis is blinded to the participant’s group assignment. Another limitation is the lack of standardisation of PFPT which can vary in quality and specific interventions between individual physiotherapists and between centres. This will be mitigated by exclusively using physiotherapists with experience and expertise in PFPT who have worked in dedicated pelvic floor centres.

Ultimately, this trial is the first prospective study to assess the utility of PFPT in the prevention of recurrent diverticulitis. It will build upon the findings from previous retrospective work, and the practical design lays the reproducible foundation for a larger prospective trial adequately powered for hypothesis testing.

### Trial status

Active recruitment is expected to begin in December 2025 and continue for 12 months or up until 40 participants have been recruited – whichever occurs first. The follow-up period for each participant is 12 months. It is therefore expected that the duration of the study will be up to 24 months (*Figure 3*). Statistical analysis will commence after all participants have completed follow-up.

**Figure 3.**
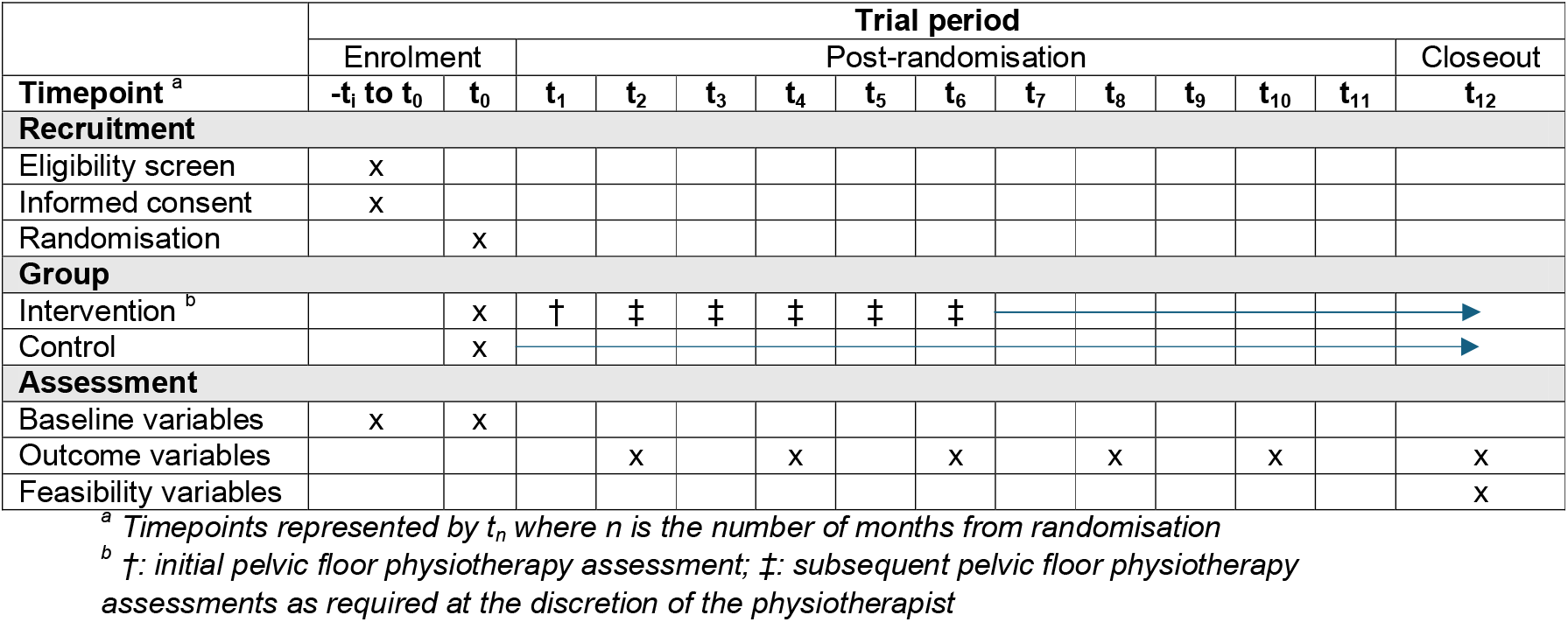
Trial schedule. ^a^ Timepoints represented by t_n_ where n is the number of months from randomisation ^b^ †: initial pelvic floor physiotherapy assessment; ‡: subsequent pelvic floor physiotherapy assessments as required at the discretion of the physiotherapist

## Data Availability

Data will be held for 15 years from the date of publication. Deidentified data will be made available on reasonable request to the corresponding author from the date of publication.

## Funding

There was no grant or financial support for this research project. There are no financial contributions to disclose.

## Conflicts of interest

None of the authors have any conflicts of interest to declare.

## Notes

### Competing Interest Statement

The authors have declared no competing interest.

### Clinical Trial

ACTRN126250009274426

### Author Declarations

Ethics approval was granted from the Metro South Health (MSH) Human Research Ethics Committee (HREC) with registration HREC/2025/QMS/120559.

